# DNA methylation patterns associated with prior tuberculosis infection in people with HIV: a pilot cross-sectional study

**DOI:** 10.1101/2025.03.19.25324254

**Authors:** Joseph Baruch Baluku, Sharon Namiiro, Daphine Kigongo, Brenda Namanda, Hakiimu Kawalya, Irene Najjingo, Waiswa Geoffrey, Nixon Niyonzima, Naghib Bogere, Edwin Nuwagira, Joshua Rhein, Nick Jones, Christian Kraef, Megan Shaughnessy, Arohi Chauhan, Immaculate Nankya, Sayoki Mfinanga, Stanton Gerson, Bruce Kirenga

## Abstract

**Background:** Mechanisms by which prior tuberculosis (TB) increases long-term risk for cancer, cardiovascular, and neurological disorders remain unclear, particularly in people with HIV (PWH). This study investigated DNA methylation (DNAm) patterns and associated pathways in PWH with and without prior TB infection.

**Methods:** DNAm was analyzed in blood samples from 30 PWH (10 with prior latent TB infection [LTBI], 10 with previous successfully treated active TB, and 10 with no TB) using the Illumina MethylationEPIC BeadChip covering over 850,000 CpG sites. Functional enrichment analyses for Gene Ontology, KEGG pathways, and gene set enrichment analysis were performed. Statistical significance was set at a false discovery rate of <0.05.

**Results:** A total of 25,084 differentially methylated CpGs (dmCpGs) were identified in the prior active TB vs. no TB comparison, corresponding to 8 differentially methylated regions (DMRs) in KCNC4-DT, GRAMD1C, ZNF44, FIGN, KCNN3, and PLA2G1B genes. In the LTBI vs. no TB comparison, 7,682 dmCpGs were observed, corresponding to 18 DMRs in SPATC1L, ZFP57, KCNN3, LRSAM1, PLEKHG5, MCF2L, BRSK2, SH3GL2, AP001468.58 and STK32C genes.

In both prior active TB vs. no TB and LTBI vs. no TB comparisons, DNAm changes were enriched in pathways related to neurogenesis, neuron differentiation, glutamatergic synapse, and neuroactive ligand-receptor interactions. The LTBI comparison showed additional enrichment in pathways related to synaptic membrane and serotonergic synapse. Cardiovascular pathways were specific to prior active TB, with significant enrichment in vascular smooth muscle contraction, arrhythmogenic right ventricular cardiomyopathy, hypertrophic cardiomyopathy, and dilated cardiomyopathy pathways.

Both TB groups showed enrichment in gene sets associated with lung, colorectal, gastric, and breast cancers. The prior active TB group demonstrated additional enrichment for prostate cancer and proteoglycans in cancer, while the LTBI group had additional enrichment for endometrial, esophageal, liver cancers, and Ewing’s sarcoma.

**Conclusion:** Prior TB infection in PWH is associated with DNAm changes in pathways related to neural function, cardiovascular health, and cancer risk suggesting epigenetic mechanisms for TB-related long-term complications.

## INTRODUCTION

Despite the improvement in diagnostic modalities and discovery of effective chemotherapeutic agents, tuberculosis (TB) is the leading cause of death from a single infectious agent, contributing 1.25 million deaths in 2023 [1]. Although the global tuberculosis treatment success for drug sensitive TB is high, long-term complications of tuberculosis contribute to significant disability and mortality even after TB treatment success [2,3]. After completion of TB treatment, previously treated patients are 3 times more likely to die than individuals without TB, and this is independent of age, sex, country income and type of TB [4] or drug resistance profile [5]. For this reason, there are growing calls for programmatic follow up of TB patients after TB cure or treatment completion [6,7]. While these calls draw attention to the several post TB pulmonary complications resulting from structural changes in the lung (such as chronic obstructive airway disease, bronchiectasis, and fibrosis) [7], cardiovascular disease (CVD) and cancer account for almost 40% of deaths after TB treatment completion, while respiratory causes contribute only 14% [4]. Evidence from cohort studies shows that pulmonary TB (PTB) increases the risk for ischemic stroke [8], acute coronary syndrome [9], myocardial infarction [10], and chronic kidney disease [11]. Additionally, PTB has been reported to increase the incidence of neurologic complications even in the absence of central nervous system tuberculosis. Dementia [12], parkinsonism [13] and depression [14] are reported in population-wide cohort studies. Finally, TB is associated with an increased risk for cancer at ten sites, including head and neck cancer, hepatobiliary cancer, Hodgkin’s lymphoma, lung cancer, gastrointestinal cancer, non-Hodgkin’s lymphoma, pancreatic cancer, leukemia, kidney and bladder cancer, and ovarian cancer [15]. Latent TB infection (LTBI) has been associated with similar risks for cancer, CVD and mental health problems [16–18].

The mechanisms underlying the long-term complications of TB are not well elucidated. Chronic inflammation and traditional CVD risk factors such as diabetes mellitus, smoking, and age have been implicated in increasing CVD risk [19,20]. Active TB and LTBI could contribute to cancer development by inducing chronic inflammation, genomic instability, and modulation of host cell signaling pathways [21]. Further, DNA methylation is thought to occur in genes and pathways involved in immune-regulation even after TB treatment. For example, this has been observed in genes involved in cytokine production (Interleukin (IL)-6), toll-like receptor signaling (TLR2), and other immune-related pathways (PI3K-AKT, MAPK, mTOR) [22]. However, these mechanisms have been demonstrated in predominantly HIV negative cohorts despite evidence that people with HIV (PWH) already experience ongoing accelerated aging driven by persistent inflammation, immune senescence, mitochondrial dysfunction, epigenetic alterations, and long term toxicities attributed to anti-retroviral therapy (ART) [23].

Our study aim was to determine whether prior TB infection is associated with DNA methylation (DNAm) patterns and pathways that could confer future risk for long term complications among PWH. By elucidating these mechanisms, the findings could inform targeted interventions to mitigate long-term complications, offering new avenues for improving the care of individuals with TB-HIV coinfection.

## METHODS

### Study design and population

In this cross-sectional study, 30 adult PWH on ART were randomly selected from three HIV clinics in Uganda. Detailed study methods are described elsewhere [24]. Participants were stratified into three groups: 10 with prior LTBI, 10 with previously treated active TB, and 10 with no history of TB infection. Previous treatment for active TB (diagnosed either by sputum GeneXpert, microscopy or urine lipoarabinomannan) was ascertained from the HIV care records and the unit TB register at the respective clinics. LTBI was defined as a positive Quantiferon (QFT)-plus assay, according to manufacturer’s instructions, in an individual without previous treatment for active TB [25]. PWH with LTBI had all completed a course of TB preventive therapy according to national guidelines prior to enrollment [26]. Prior TB was defined as either LTBI or previous treated active TB. PWH with no TB had never been treated for active TB and had a negative QFT-plus assay.

### Sample collection and processing

Whole blood samples (5 ml) were collected from each participant using EDTA vacutainers. Genomic DNA was extracted from the blood using the QIAamp DNA Blood Mini Kit (QIAGEN, Hilden, Germany) according to the manufacturer’s instructions. DNA quantity was measured using Qubit 4 fluorometer (ThermoFisher Scientific, USA), following the manufacturer’s instructions. For the methylation assay, 500 ng of DNA was used. DNA methylation (DNAm) levels were assayed using the Illumina Infinium MethylationEPIC v2.0 BeadChip array and the iScan system (Illumina, Inc., San Diego, CA, USA), in accordance with the manufacturer’s instructions, generating IDAT files for downstream bioinformatics analysis to assess methylation levels.

### Data processing and analysis

The raw data files obtained from the Illumina MethylationEPIC BeadChip were preprocessed and analyzed using the minfi package in the R statistical software environment. The minfi package is a comprehensive toolkit specifically designed for the analysis of Illumina Infinium methylation arrays [27]. The preprocessing steps involved an initial quality assessment of the raw data using various metrics, including visualization of beta value distributions and evaluation of control probes to ensure optimal bisulfite conversion efficiency. Subsequently, background correction and quantile normalization were performed using the preprocess Quantile function to minimize technical variation and ensure consistent methylation value distributions across samples. To ensure data quality, probes with a detection p-value greater than 0.01 were removed, and probes located on sex chromosomes (X and Y) were excluded to avoid confounding due to sex differences. Additionally, probes containing single nucleotide polymorphisms (SNPs) were removed using the dropLociWithSnps function to prevent methylation differences caused by genetic variation.

Blood cell composition was estimated using the estimateCellCounts2 function, referencing the FlowSorted.CordBloodCombined.450k dataset and utilizing the IDOLOptimizedCpGsCordBlood marker set. The cell types assessed included CD4+ T cells, CD8+ T cells, B cells, monocytes, natural killer (NK) cells, granulocytes, and nucleated red blood cells. The consistent proportion of blood cell types across samples, as detailed in **Supplementary Table 1**, indicated the absence of biological bias in the experimental setup. This consistency is important because the selected Horvath’s methylation clock, used for age estimation, incorporates various cell types, including blood cells.

Methylation beta values were also used to estimate epigenetic age for each individual using Horvath, a first-generation DNAm epigenetic clock [28]. Age acceleration represented the extent to which the biological age of an individual exceeds their chronological age at the time of measurement. Age estimates were determined using dnaMethyAge package version 0.2.0 and the measure for age acceleration was defined as residuals yielded from regressing epigenetic age on chronological age from which root mean squared deviation (RMSD) and mean absolute deviation (MAD) were computable.

### Identification of differentially methylated CpGs and regions

Differentially methylated CpGs (dmCpGs) and regions (DMRs) were identified using the dmpFinder function in minfi, which applies a linear model to compare methylation levels while adjusting for age and sex. This function performed pairwise comparisons between the three groups (LTBI, active TB, and no TB) to identify CpG sites that exhibit significant differences in methylation levels. Significant regions were defined by β-value differences >0.2 and permutation testing (B=0). The dmpFinder function utilizes statistical tests to assess the significance of methylation differences, adjusting for multiple comparisons to control the false discovery rate (FDR). An FDR <0.05 was considered statistically significant. The Comparisons included: 1. Previous Active TB vs. no TB; 2. Prior LTBI TB vs. no TB; 3. Prior TB infection (both LTBI and previous active TB) vs. no TB; and 4. Previous active TB vs. LTBI.

### Pathway Analysis

Pathway and gene ontology (GO) enrichment analyses were conducted using the missMethyl package to identify biological pathways associated with significant DMRs. These included GO enrichment analysis [29] using the gometh function and pathway analysis through the gsameth function, leveraging gene sets from the Kyoto Encyclopedia of Genes and Genomes (KEGG) [30], Molecular Signatures Database (MSigDB) [31] and PANTHER version 16 database [32]. All statistical analyses were carried out using R version 4.3.1.

## RESULTS

### Characteristics of study participants of PWH with and without prior TB

Of 30 participants, 17 (57%) were female and the median (interquartile range, IQR) age was 46.5 (40.0 – 50.0) years. All participants were on ART and had achieved viral load suppression (viral load <1,000 copies/ml). **Table 1** shows characteristics of the participants.

**Table 1:**
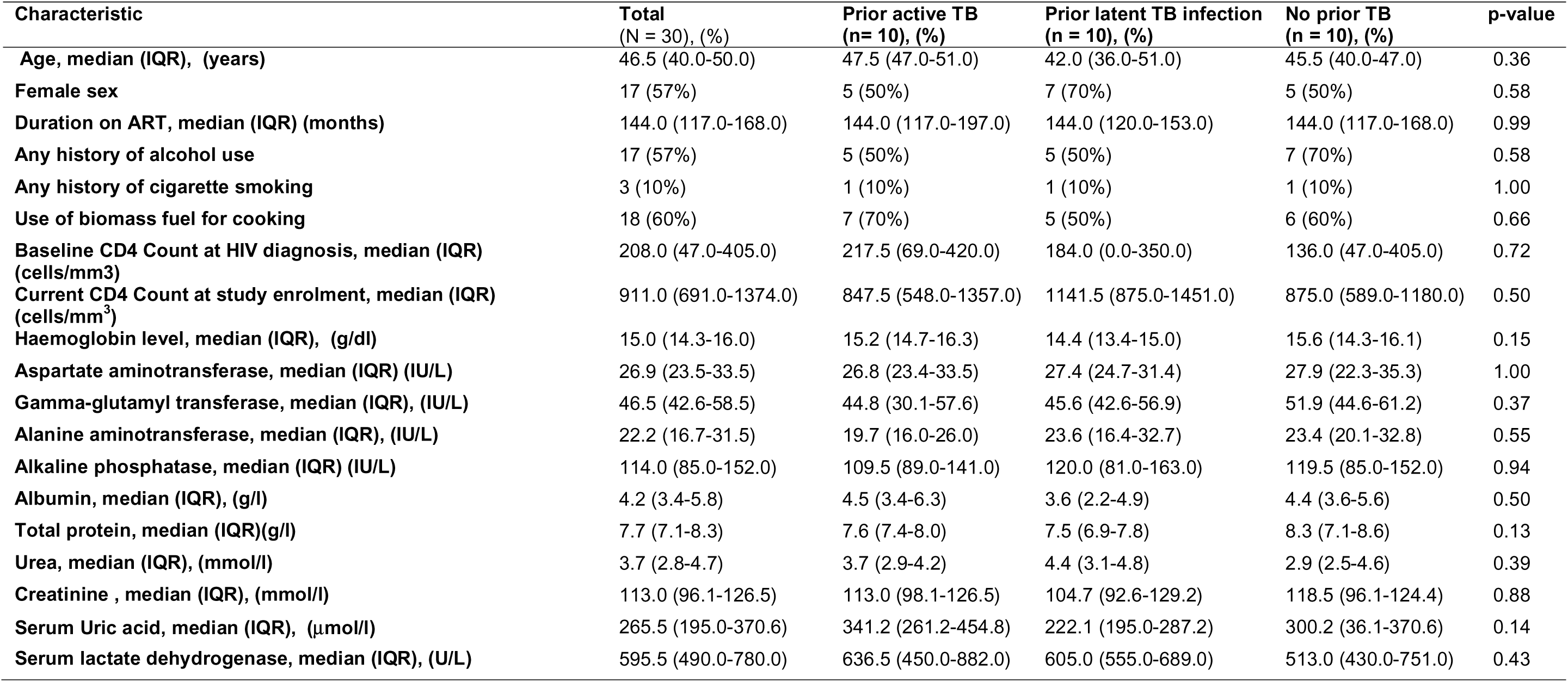
Characteristics of study participants.

| Characteristic | Total<br>(N = 30), (%) | Prior active TB<br>(n= 10), (%) | Prior latent TB infection<br>(n = 10), (%) | No prior TB<br>(n = 10), (%) | p-value |
| --- | --- | --- | --- | --- | --- |
| Age, median (IQR), (years) | 46.5 (40.0-50.0) | 47.5 (47.0-51.0) | 42.0 (36.0-51.0) | 45.5 (40.0-47.0) | 0.36 |
| Female sex | 17 (57%) | 5 (50%) | 7 (70%) | 5 (50%) | 0.58 |
| Duration on ART, median (IQR) (months) | 144.0 (117.0-168.0) | 144.0 (117.0-197.0) | 144.0 (120.0-153.0) | 144.0 (117.0-168.0) | 0.99 |
| Any history of alcohol use | 17 (57%) | 5 (50%) | 5 (50%) | 7 (70%) | 0.58 |
| Any history of cigarette smoking | 3 (10%) | 1 (10%) | 1 (10%) | 1 (10%) | 1.00 |
| Use of biomass fuel for cooking | 18 (60%) | 7 (70%) | 5 (50%) | 6 (60%) | 0.66 |
| Baseline CD4 Count at HIV diagnosis, median (IQR)<br>(cells/mm<sup>3</sup>) | 208.0 (47.0-405.0) | 217.5 (69.0-420.0) | 184.0 (0.0-350.0) | 136.0 (47.0-405.0) | 0.72 |
| Current CD4 Count at study enrolment, median (IQR)<br>(cells/mm<sup>3</sup>) | 911.0 (691.0-1374.0) | 847.5 (548.0-1357.0) | 1141.5 (875.0-1451.0) | 875.0 (589.0-1180.0) | 0.50 |
| Haemoglobin level, median (IQR), (g/dl) | 15.0 (14.3-16.0) | 15.2 (14.7-16.3) | 14.4 (13.4-15.0) | 15.6 (14.3-16.1) | 0.15 |
| Aspartate aminotransferase, median (IQR) (IU/L) | 26.9 (23.5-33.5) | 26.8 (23.4-33.5) | 27.4 (24.7-31.4) | 27.9 (22.3-35.3) | 1.00 |
| Gamma-glutamyl transferase, median (IQR), (IU/L) | 46.5 (42.6-58.5) | 44.8 (30.1-57.6) | 45.6 (42.6-56.9) | 51.9 (44.6-61.2) | 0.37 |
| Alanine aminotransferase, median (IQR), (IU/L) | 22.2 (16.7-31.5) | 19.7 (16.0-26.0) | 23.6 (16.4-32.7) | 23.4 (20.1-32.8) | 0.55 |
| Alkaline phosphatase, median (IQR) (IU/L) | 114.0 (85.0-152.0) | 109.5 (89.0-141.0) | 120.0 (81.0-163.0) | 119.5 (85.0-152.0) | 0.94 |
| Albumin, median (IQR), (g/l) | 4.2 (3.4-5.8) | 4.5 (3.4-6.3) | 3.6 (2.2-4.9) | 4.4 (3.6-5.6) | 0.50 |
| Total protein, median (IQR)(g/l) | 7.7 (7.1-8.3) | 7.6 (7.4-8.0) | 7.5 (6.9-7.8) | 8.3 (7.1-8.6) | 0.13 |
| Urea, median (IQR), (mmol/l) | 3.7 (2.8-4.7) | 3.7 (2.9-4.2) | 4.4 (3.1-4.8) | 2.9 (2.5-4.6) | 0.39 |
| Creatinine , median (IQR), (mmol/l) | 113.0 (96.1-126.5) | 113.0 (98.1-126.5) | 104.7 (92.6-129.2) | 118.5 (96.1-124.4) | 0.88 |
| Serum Uric acid, median (IQR), (μmol/l) | 265.5 (195.0-370.6) | 341.2 (261.2-454.8) | 222.1 (195.0-287.2) | 300.2 (36.1-370.6) | 0.14 |
| Serum lactate dehydrogenase, median (IQR), (U/L) | 595.5 (490.0-780.0) | 636.5 (450.0-882.0) | 605.0 (555.0-689.0) | 513.0 (430.0-751.0) | 0.43 |

### Chronological and epigenetic age of i PWH with and without prior TB

The median chronological and epigenetic ages for the whole study population were 46.5 and 51.5 years, respectively. When stratified by group, the median chronological and epigenetic ages were as follows: previous active TB (47.5 vs. 53.2 years), LTBI (42.0 vs. 47.4 years), and no TB groups (45.5 vs. 49.6 years). The comparison of epigenetic and chronological age between prior TB infection (both LTBI and active) and those without TB is shown in **figure 1**. The results demonstrate an overall trend toward accelerated aging among PWH that was more pronounced in people with prior TB infection (both LTBI and active TB).

**Figure 1:**
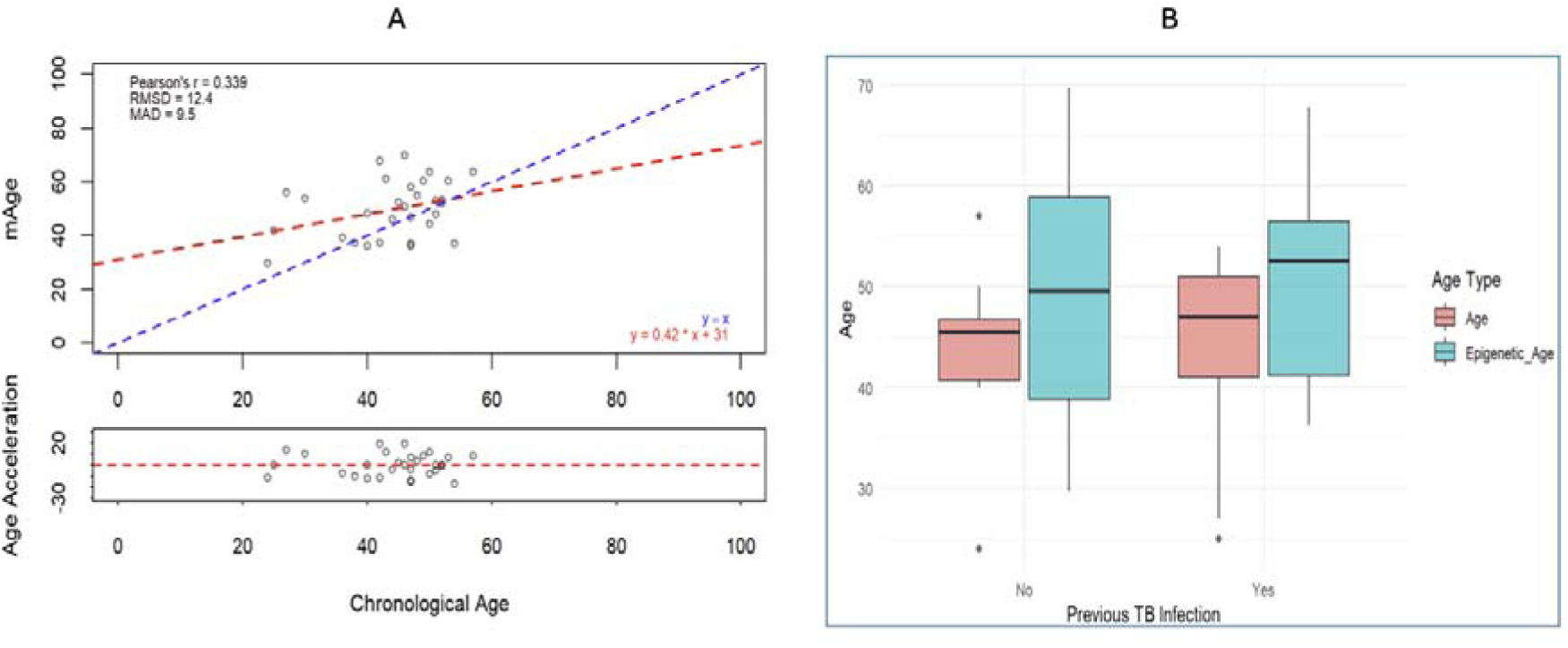
Epigenetic Age (DNAm) Compared to Chronological Age in Individuals with and without Prior Tuberculosis Infection. (A) Scatter plot of DNAm age(y-axis) versus chronological age (x-axis) estimated using Horvath’s 2018 methylation clock. The blue dashed line represents perfect agreement between DNAm age and chronological age, while the red dashed line represents the regression line derived from the actual data. Most samples are above the blue line, indicating an overestimation of DNAm age. The bottom panel shows age acceleration values (DNAm age minus chronological age residuals), with positive values indicating accelerated aging (DNAm age > chronological age) and negative values indicating decelerated aging (DNAm age < chronological age). The results demonstrate a slight overall trend toward accelerated aging. (B) Box plot comparing DNAm age (Epigenetic_Age) and chronological age (Age) between individuals with previous TB infection and those without TB infection. DNAm age is higher than chronological age in both groups, and individuals with a history of TB show slightly higher DNAm age compared to the group without prior TB. This suggests potential epigenetic age acceleration associated with prior TB infection.

### DNA methylation patterns in PWH with and without prior TB

A total of 25,084 differentially methylated CpGs (dmCpGs) were identified in the comparison between prior active TB vs. no TB groups (**Table 2 and Figure 2: A1&A2**). These dmCpGs corresponded to 8 differentially methylated regions (DMRs) associated with the following genes: KCNC4-DT (hypomethylation), FIGN (hypomethylation), MS4A4A (hypomethylation), GRAMD1C (hypermethylation), ZNF44 (hypermethylation), KCNN3 (hypermethylation), and PLA2G1B (hypermethylation).

**Figure 2:**
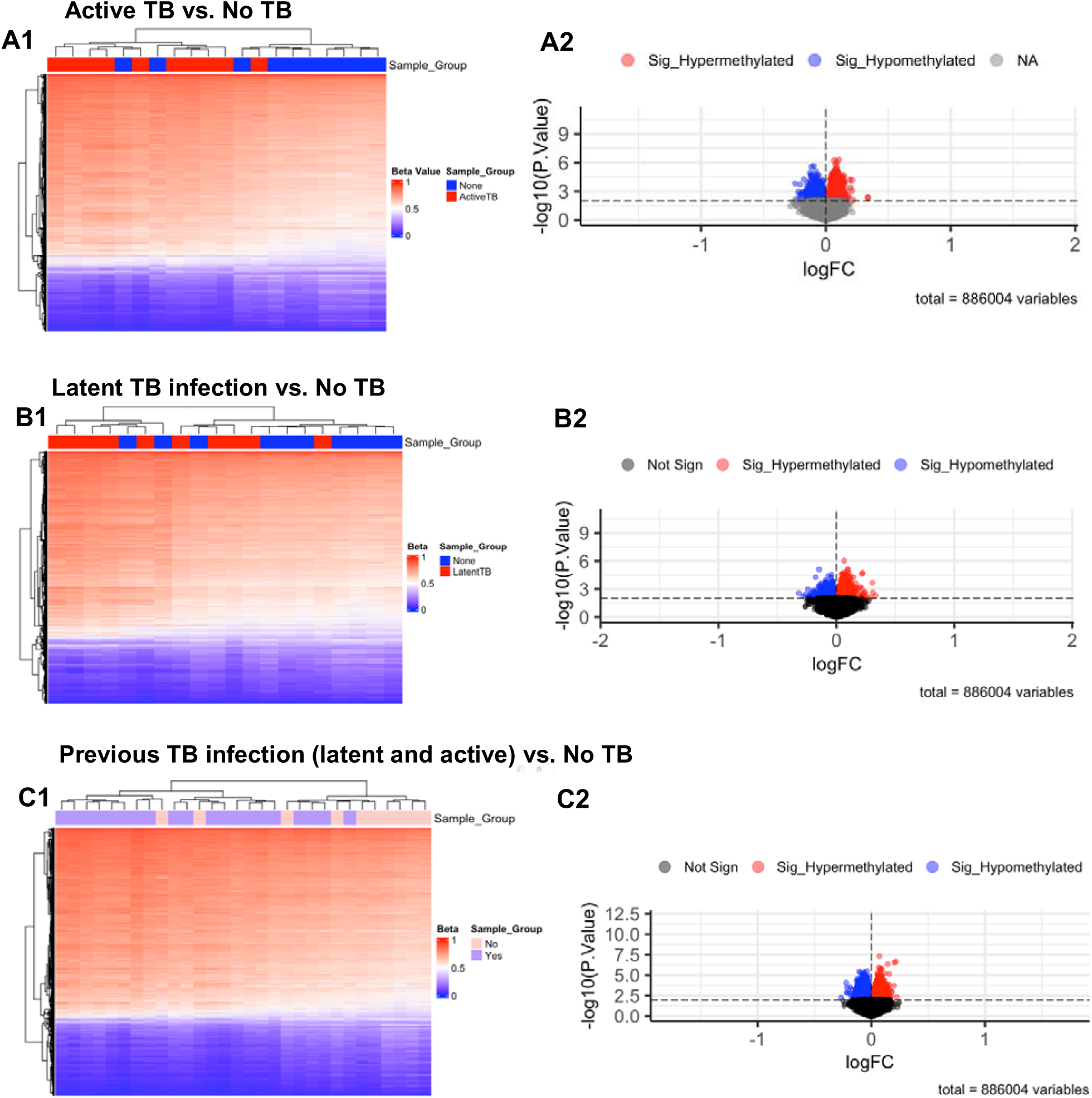
Heatmaps A1, B1, and C1 derived from beta values of the differently methylated CpG sites identified in pairwise comparison across different groups with stringency criteria of adjusted P < 0.01 and mean methylation difference >0.2. Dendrogram shows separation based on groups. Enhanced Volcano plots A2, B2 and C2 of the differentially methylated CpG-sites separating different groups. The x-axis is the log fold change and the y-axis is the negative log10 of p-value with the cut off p-value of 0.05 shown with dash-dotted horizontal line. Blue CpGs indicate hypomethylated while red CpGs are hypermethylated.

**Table 2:** DNA methylation patterns among people with HIV with and without prior TB infection.

| Chromosome | Start | End | SNP | Gene | Methylation Value | Area |
| --- | --- | --- | --- | --- | --- | --- |
| **Active TB Vs. No TB** |  |  |  |  |  |  |
| Chr1 | 110209635 | 110209685 | rs1280981579 | KCNC4-DT | -0.2273238 | 0.4546476 |
| Chr3 | 113890077 | 113890077 | rs1026956222 | GRAMD1C | 0.3387863 | 0.3387863 |
| Chr19 | 12230577 | 12230577 | rs1412969463 | ZNF44 | 0.3342976 | 0.3342976 |
| Chr2 | 163733762 | 163733762 | rs867364910 | FIGN | -0.2206179 | 0.2206179 |
| Chr1 | 154867337 | 154867337 | rs1015124254 | KCNN3 | 0.2084786 | 0.2084786 |
| Chr12 | 120326012 | 120326012 | rs746790393 | PLA2G1B | 0.2078026 | 0.2078026 |
| Chr3 | 46665072 | 46665072 | rs1309260839 | Non-exon region | -0.2070116 | 0.2070116 |
| Chr11 | 60251763 | 60251763 | rs112829924 | MS4A4A | -0.2055199 | 0.2055199 |
| **Latent TB vs. No TB** |  |  |  |  |  |  |
| chr21 | 46161128 | 46161644 | - | SPATC1L | -0.2251591 | 0.6754773 |
| chr6 | 29680827 | 29681124 | - | ZFP57 | -0.2725237 | 0.5450474 |
| chr21 | 46184138 | 46184252 | - | SPATC1L | 0.272069 | 0.544138 |
| chr1 | 154867337 | 154867343 | - | KCNN3 | 0.2201583 | 0.4403155 |
| chr9 | 127479094 | 127479094 | rs983018719 | LRSAM1 | 0.3328147 | 0.3328147 |
| chr1 | 6491645 | 6491645 | rs1042531521 | PLEKHG5 | -0.3182199 | 0.3182199 |
| chr21 | 46185260 | 46185260 | rs1458119335 | AP001468.58 | 0.304427 | 0.3044274 |
| chr13 | 113176555 | 113176555 | rs1011009098 | Non-exon region | 0.2545671 | 0.2545671 |
| Chr17 | 16997705 | 16997705 | rs756899690 | Non-exon region | 0.2430883 | 0.2430883 |
| chr13 | 112980664 | 112980664 | rs1040730563 | MCF2L | -0.2410457 | 0.2410457 |
| chr2 | 127695686 | 127695686 | rs996351088 | Non-exon region | 0.2223182 | 0.2223182 |
| chr11 | 1391915 | 1391915 | rs1298386467 | BRSK2 | -0.2193803 | 0.2193803 |
| chr4 | 183349316 | 183349316 | rs751119789 | Non-exon region | 0.2173798 | 0.2173798 |
| chr22 | 47918625 | 47918625 | rs536912378 | CTA-280A3.2 | 0.2166322 | 0.2166322 |
| chr9 | 17612106 | 17612106 | rs750183679 | SH3GL2 | 0.2113893 | 0.2113893 |
| chr10 | 132231205 | 132231205 | rs1163463011 | STK32C | -0.2069165 | 0.2069165 |
| chr5 | 151613206 | 151613206 | rs956799603 | Non-exon region | -0.2026999 | 0.2026999 |
| chr11 | 1435923 | 1435923 | rs551352724 | BRSK2 | 0.2009539 | 0.2009539 |
| **Previous TB infection (both active and latent TB) Vs. No TB** |  |  |  |  |  |  |
| chr1 | 154867337 | 154867433 | rs1015124254 | KCNN3 | 0.2100413 | 0.4200826 |
| chr1 | 6491645 | 6491645 | rs1042531521 | PLEKHG5 | -0.2672408 | 0.2672408 |
| chr13 | 113176555 | 113176555 | rs1011009098 | Non-exon region | 0.2242132 | 0.2242132 |
| chr1 | 110209685 | 110209685 | rs188603653 | KCNC4-DT | -0.2226747 | 0.2226747 |
| chr13 | 112980664 | 112980664 | rs1040730563 | MCF2L | -0.2195602 | 0.2195602 |
| chr10 | 59654071 | 59654071 | rs761027162 | SLC16A9 | -0.2140531 | 0.2140531 |
| chr11 | 1391915 | 1391915 | rs1298386467 | BRSK2 | -0.2074399 | 0.2074399 |
| chr4 | 183349316 | 183349316 | rs751119789 | Non-exon region | 0.2006364 | 0.2006364 |
| **Latent TB Vs. active TB** |  |  |  |  |  |  |
| chr19 | 50051194 | 50051194 |  | ZNF473,<br>FLJ26850 | -0.2943314 | 0.2943314 |
| chr2 | 30446891 | 30446891 | rs1681268169 | Non-exon region | -0.2796443 | 0.2796443 |
| chr2 | 30446893 | 30446893 | rs1327445118 | Non-exon region | -0.2789843 | 0.2789843 |
| chr21 | 46185260 | 46185260 | rs1458119335 | AP001468.58 | -0.2477524 | 0.2477524 |
| chr16 | 68070553 | 68070553 | rs959716619 | DUS2 | -0.2143316 | 0.2143316 |
| chr2 | 201666059 | 201666059 | rs918060583 | MPP4 | 0.2120617 | 0.2120617 |

In the comparison between LTBI vs. no TB, 7,682 dmCpGs were observed (**Table 2 and Figure 2: B1&2**). These were associated with 18 DMRs, including genes such as ZFP57 (hypomethylation), PLEKHG5 (hypomethylation), STK32C (hypomethylation), MCF2L (hypomethylation), KCNN3 (hypermethylation), LRSAM1 (hypermethylation), AP001468.58 (hypermethylation), CTA-280A3.2 or EPIC1-LOC124905150 (hypermethylation), SH3GL2 (hypermethylation) as well as SPATC1L and BRSK2 gene regions with both hypo and hypermethylated probes.

Overall, 38,558 dmCpGs were identified when comparing individuals with any history of TB infection (both active and latent TB) to the control group (**Table 2 and Figure 2: C1&2**). These dmCpGs corresponded to 8 DMRs associated with genes such as KCNC4-DT (hypomethylation), PLEKHG5 (hypomethylation), SLC16A9 (hypomethylation), BRSK2 (hypomethylation), MCF2L (hypomethylation) and KCNN3 (hypermethylation).

### Enriched pathways associated with DNAm patterns in PWH with and without prior TB

In both prior active TB vs. no TB and prior LTBI vs. no TB comparisons, DNAm changes were enriched in pathways related to neurogenesis and neuron differentiation. Comparing groups exposed to prior TB (both active and LTBI) to controls revealed enrichment in pathways related to arrhythmogenic right ventricular cardiomyopathy, proteoglycans in cancer, neuroactive ligand-receptor interactions, axonal guidance, glutamatergic synapses, lung cancer, and breast cancer (**Figure 3A**).

**Figure 3:**
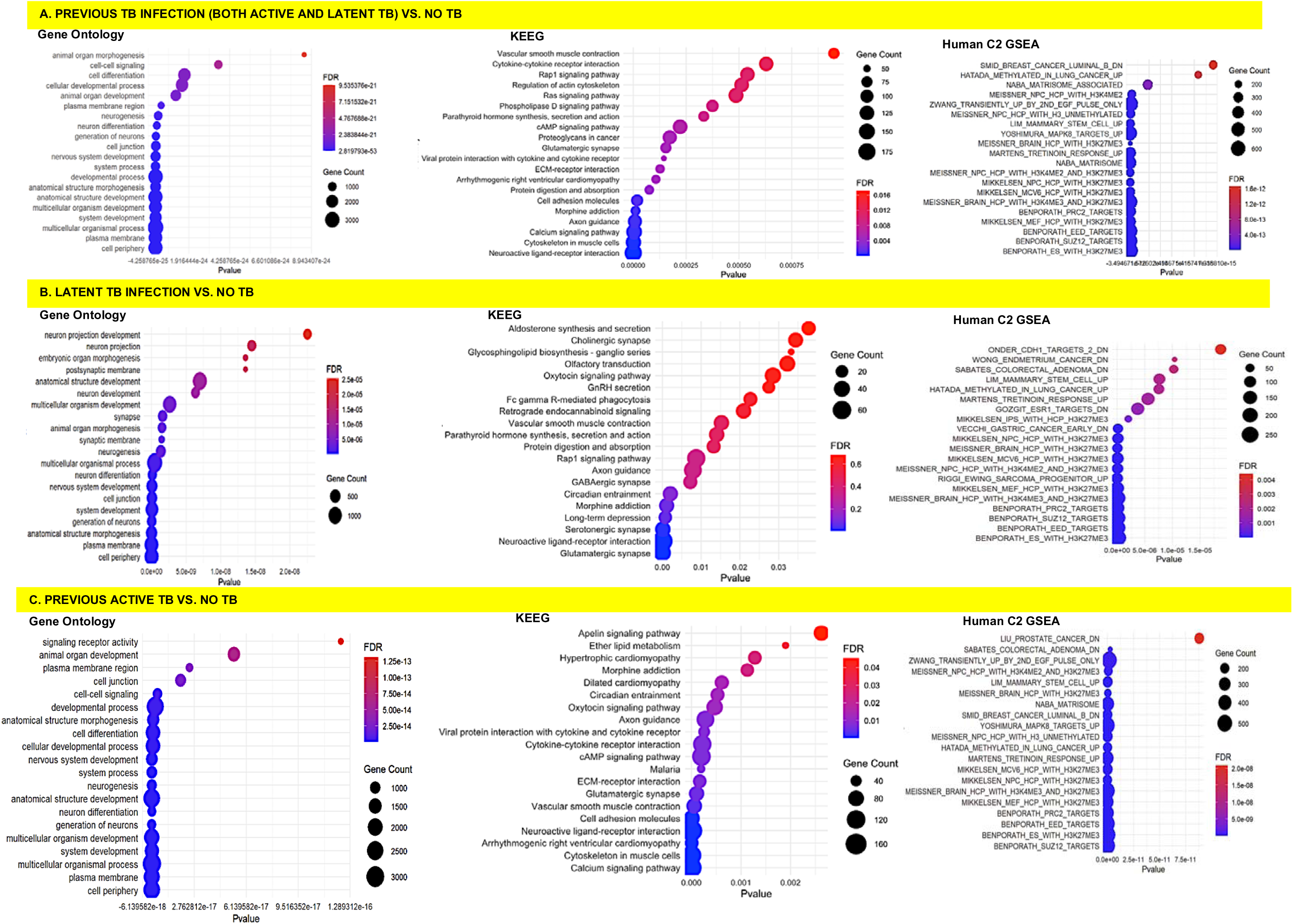
Pathway enrichment analysis for association of prior TB and DNA methylation. In each panel, the x-axis represents the p-value for pathway enrichment (adjusted for false discovery rate [FDR]), while the y-axis lists the enriched pathways. The size of the dots indicates the number of genes involved in each pathway, and the color gradient represents the FDR, with red indicating higher significance (lower FDR)

LTBI showed additional enrichment in pathways related to glutamatergic and serotonergic synapses, neuroactive ligand-receptor interactions (**Figure 3B**). Cardiovascular pathways were specific to prior active TB, with enrichment in vascular smooth muscle contraction, arrhythmogenic right ventricular cardiomyopathy, hypertrophic cardiomyopathy, and dilated cardiomyopathy pathways (**Figure 3C**).

Both pairwise comparisons (prior active TB vs. no TB and prior LTBI vs. no TB comparisons) showed enrichment in gene sets associated with lung, colorectal, gastric, and breast cancers. The prior active TB group demonstrated additional enrichment for prostate cancer and proteoglycans in cancer, while the LTBI group had additional enrichment for endometrial, esophageal, liver cancers, and Ewing’s sarcoma.

Analysis using the PANTHER database consistently showed enrichment in the Wnt signaling pathway, with 34 associated genes in the active TV vs no TB comparison and 17 genes in the latent TB vs. no TB comparison. Pathways involved in inflammation mediated by chemokine and cytokine signaling, integrin signaling, and angiogenesis were also enriched (**Table 3**).

**Table 3:**
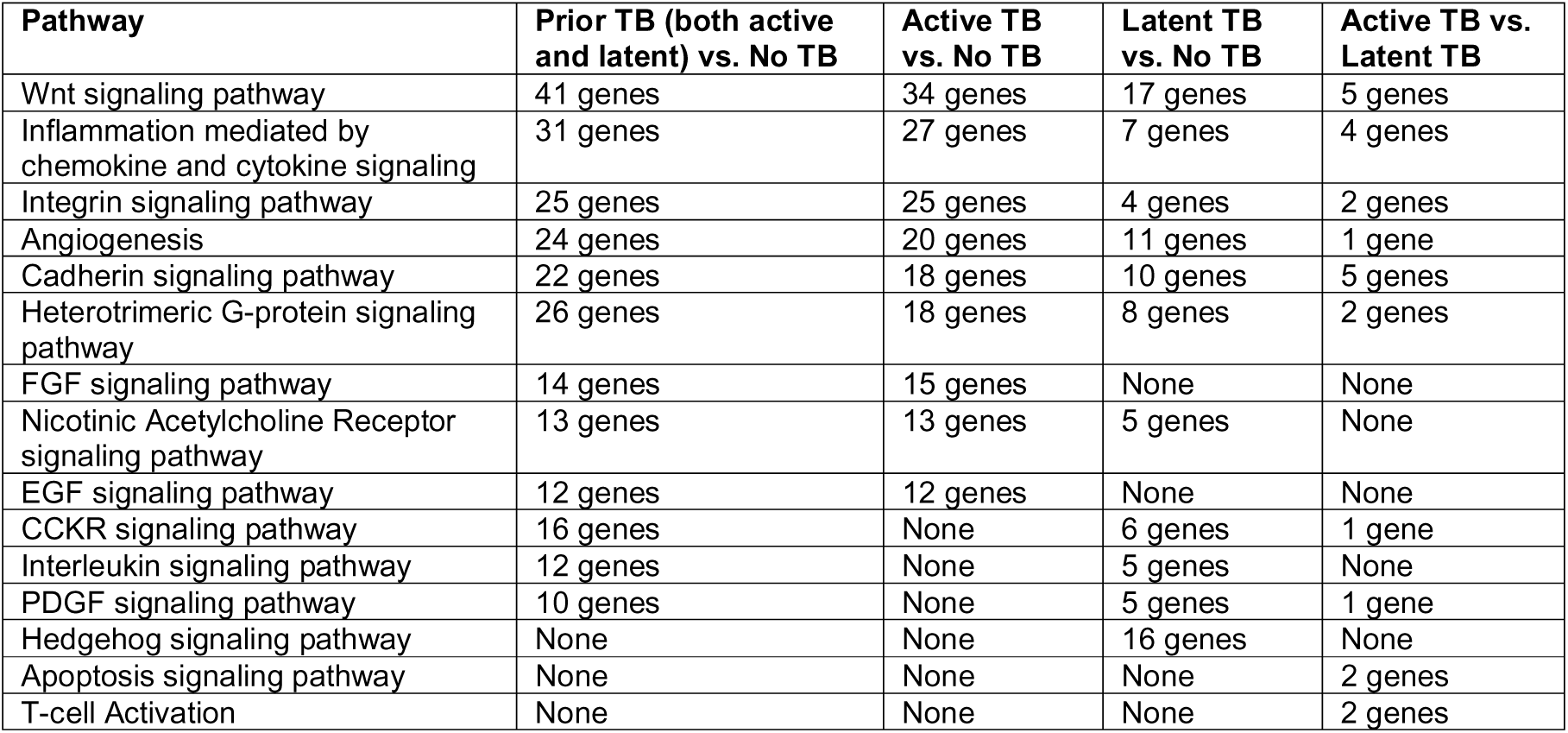
Pathway enrichment in PWH with and without prior TB from the PANTHER database.

| Pathway | Prior TB (both active and latent) vs. No TB | Active TB vs. No TB | Latent TB vs. No TB | Active TB vs. Latent TB |
| --- | --- | --- | --- | --- |
| Wnt signaling pathway | 41 genes | 34 genes | 17 genes | 5 genes |
| Inflammation mediated by chemokine and cytokine signaling | 31 genes | 27 genes | 7 genes | 4 genes |
| Integrin signaling pathway | 25 genes | 25 genes | 4 genes | 2 genes |
| Angiogenesis | 24 genes | 20 genes | 11 genes | 1 gene |
| Cadherin signaling pathway | 22 genes | 18 genes | 10 genes | 5 genes |
| Heterotrimeric G-protein signaling pathway | 26 genes | 18 genes | 8 genes | 2 genes |
| FGF signaling pathway | 14 genes | 15 genes | None | None |
| Nicotinic Acetylcholine Receptor signaling pathway | 13 genes | 13 genes | 5 genes | None |
| EGF signaling pathway | 12 genes | 12 genes | None | None |
| CCKR signaling pathway | 16 genes | None | 6 genes | 1 gene |
| Interleukin signaling pathway | 12 genes | None | 5 genes | None |
| PDGF signaling pathway | 10 genes | None | 5 genes | 1 gene |
| Hedgehog signaling pathway | None | None | 16 genes | None |
| Apoptosis signaling pathway | None | None | None | 2 genes |
| T-cell Activation | None | None | None | 2 genes |

## DISCUSSION

In this study, we investigated DNAm patterns associated with prior TB infection in individuals with HIV. Our findings reveal that both prior active TB and LTBI are associated with distinct DNAm changes, particularly in pathways related to neural function and development, cardiovascular health, and cancer risk. In addition, there was a trend suggesting that prior TB accelerates aging among PWH. These observations suggest potential mechanisms through which TB infection may contribute to long-term complications in individuals with HIV.

Our study identified numerous dmCpGs and DMRs associated with prior TB infection. Notably, both prior active TB and LTBI were associated with DNAm changes in pathways related to neurogenesis and neuron differentiation. LTBI showed additional enrichment in pathways related to glutamatergic and serotonergic synapses, and neuroactive ligand-receptor interactions, potentially indicating an impact on synaptic plasticity and neurotransmission mediated through the Wnt signaling pathway and inflammation pathways as shown by our enrichment analysis from the PANTHER database [33]. Interestingly there was a trend in enrichment of pathways related to long-term depression as well (p=7.57×10^−4^, FDR = 0.068). These findings suggest that prior TB infection, even in the absence of active disease, may have long-term consequences for neurocognitive function in PWH [34].

Furthermore, we observed distinct DNAm changes related to cardiovascular health. Prior active TB was specifically associated with enrichment in pathways related to vascular smooth muscle contraction, arrhythmogenic right ventricular cardiomyopathy, hypertrophic cardiomyopathy, and dilated cardiomyopathy. It is unclear whether this is through TB’s modification of traditional CVD risk factors or a direct effect of TB on the myocardium [19,20]. However, our previous work among PWH with prior active TB suggests that prior TB is associated with normal body mass index, lower prevalence of dyslipidemia and no significant effect on hypertension risk [35]. Therefore, more studies are needed to understand mechanisms by which prior pulmonary TB affects the cardiac rhythm, myocardium and myocardial contractility.

Our study also revealed DNAm changes in pathways associated with various cancers. Both prior active TB and LTBI showed enrichment in gene sets associated with lung, colorectal, gastric, and breast cancers, suggesting a potential link between prior TB infection and an increased risk of these cancers. Additionally, prior active TB demonstrated enrichment for prostate cancer and proteoglycans in cancer, while LTBI showed enrichment for endometrial, esophageal, liver cancers, and Ewing’s sarcoma. These findings suggest that prior TB infection may influence cancer risk in individuals with HIV through epigenetic modifications. This is an agreement with epidemiological studies that have demonstrated that TB is associated with higher risk for incident lung, gastrointestinal, and tumors of the reproductive system tumors [15,36] Moreover, the prominent enrichment in the gene sets associated with embryonic stem cell identity: BENPORATH_ES_WITH_H3K27ME3, BENPORATH_EED_TARGETS, and BENPORATH_SUZ12_TARGETS suggests potential risk for the development of aggressive, poorly differentiated, and estrogen receptor-negative tumors [37]. The association between prior TB and advanced cancer types needs to be demonstrated in epidemiological studies.

The mechanisms underlying the observed DNAm changes and their specific contributions to long-term complications warrant further investigation by larger studies. It is possible that chronic inflammation and immune dysregulation associated with TB infection may lead to persistent epigenetic modifications, even after the resolution of active disease [22]. Additionally, the interaction between TB infection, HIV infection, and antiretroviral therapy may further contribute to epigenetic changes and long-term complications.

Our study has several limitations. The cross-sectional design limits our ability to establish causal relationships between prior TB infection and DNAm changes. The relatively small sample size may have limited our power to detect additional significant associations.

Furthermore, our study focused on peripheral blood DNAm, which may not fully reflect epigenetic changes in other tissues relevant to long-term complications.

## CONCLUSION

The findings of this study have several critical implications for clinical care, and public health. First, the identification of DNAm patterns associated with prior TB infection highlights the potential for developing predictive biomarkers. These biomarkers could enable early risk stratification in PWH, allowing targeted interventions to mitigate long-term complications such as CVD, neurocognitive disorders, and cancers.

Second, integrating epigenetic profiling into routine TB-HIV care could pave the way for personalized medicine. Tailored follow-up protocols based on DNAm profiles could optimize screening and preventive care, particularly in resource-limited settings where resource constraints demand efficient allocation of healthcare resources. Nonetheless, further research is needed to elucidate the specific mechanisms underlying these epigenetic modifications and to explore potential interventions to mitigate the long-term risks associated with prior TB infection in individuals with HIV.

## Data Availability

Datasets used in this study are available from the corresponding author upon reasonable request.

## LIST OF ABBREVIATIONS

ART: Antiretroviral Therapy
CVD: Cardiovascular Disease
DMR: Differentially Methylated Region
DNAm: DNA Methylation
dmCpGs: Differentially Methylated CpG Sites
FDR: False Discovery Rate
HIV: Human Immunodeficiency Virus
KEGG: Kyoto Encyclopedia of Genes and Genomes
LTBI: Latent Tuberculosis Infection
MSigDB: Molecular Signatures Database
NK: Natural Killer (cells)
PANTHER: Protein Analysis Through Evolutionary Relationships
PWH: People with HIV
QFT: QuantiFERON
RMSD: Root Mean Squared Deviation
TB: Tuberculosis

## DECLARATIONS

### Ethics approval and consent to participate

Study participants provided written informed consent before study measurements were undertaken. The study protocol was approved by the Mildmay Uganda Research and Ethics Committee (#REC REF MUREC-107-2022). Further, the Uganda National Council of Science and Technology provided additional approval as required by the guidelines for conducting research in Uganda (HS2328ES). All methods were performed in accordance with the relevant guidelines and regulations.

### Consent for publication

Not applicable

### Availability of data and materials

Datasets used in this study are available from the corresponding author upon reasonable request.

### Authors’ contributions

JBB – conceptualization, methodology, investigation, data accrual, formal analysis, interpretation of results, drafting manuscript, editing manuscript, final approval

SN - investigation, data accrual, interpretation of results, drafting manuscript, editing manuscript, final approval

DK - investigation, data accrual, interpretation of results, drafting manuscript, editing manuscript, final approval

BN - investigation, data accrual, interpretation of results, drafting manuscript, editing manuscript, final approval

HK – methodology, investigation, data accrual, formal analysis, interpretation of results, drafting manuscript, editing manuscript, final approval

IJ - interpretation of results, drafting manuscript, editing manuscript, final approval

WG – methodology, investigation, formal analysis, interpretation of results, drafting manuscript, editing manuscript, final approval

NN - investigation, interpretation of results, drafting manuscript, editing manuscript, final approval

NB - investigation, interpretation of results, drafting manuscript, editing manuscript, final approval

EW - investigation, data accrual, interpretation of results, drafting manuscript, editing manuscript, final approval

JR - investigation, interpretation of results, drafting manuscript, editing manuscript, final approval

NJ - investigation, interpretation of results, drafting manuscript, editing manuscript, final approval

CK - investigation, interpretation of results, drafting manuscript, editing manuscript, final approval

MS - investigation, interpretation of results, drafting manuscript, editing manuscript, final approval

AC - investigation, interpretation of results, drafting manuscript, editing manuscript, final approval

IN – methodology, investigation, data accrual, formal analysis, interpretation of results, drafting manuscript, editing manuscript, final approval

SM - investigation, interpretation of results, drafting manuscript, editing manuscript, final approval

SG - investigation, interpretation of results, drafting manuscript, editing manuscript, final approval

BK - investigation, dinterpretation of results, drafting manuscript, editing manuscript, final approval

## Acknowledgements

None

---

**Supplementary Table:**
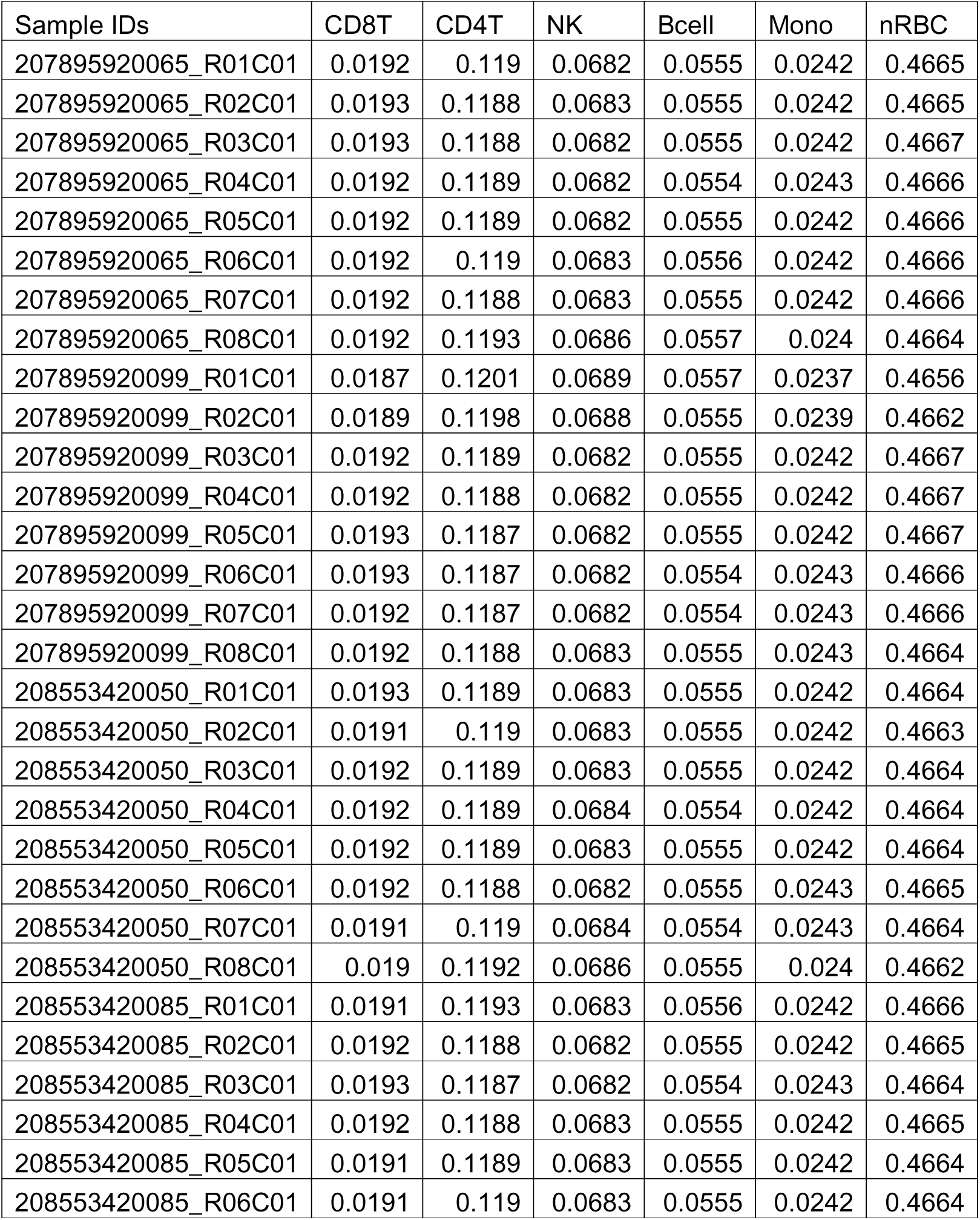
Propotion of blood Cell counts in the different study samples.

