## Supplementary material for "DNA methylation patterns associated with prior tuberculosis infection in people with HIV: a pilot cross-sectional study"

**Supplementary Table: Proportion of blood Cell counts in the different study samples**

| Sample IDs | CD8T | CD4T | NK | Bcell | Mono | nRBC |
| --- | --- | --- | --- | --- | --- | --- |
| 207895920065_R01C01 | 0.0192 | 0.119 | 0.0682 | 0.0555 | 0.0242 | 0.4665 |
| 207895920065_R02C01 | 0.0193 | 0.1188 | 0.0683 | 0.0555 | 0.0242 | 0.4665 |
| 207895920065_R03C01 | 0.0193 | 0.1188 | 0.0682 | 0.0555 | 0.0242 | 0.4667 |
| 207895920065_R04C01 | 0.0192 | 0.1189 | 0.0682 | 0.0554 | 0.0243 | 0.4666 |
| 207895920065_R05C01 | 0.0192 | 0.1189 | 0.0682 | 0.0555 | 0.0242 | 0.4666 |
| 207895920065_R06C01 | 0.0192 | 0.119 | 0.0683 | 0.0556 | 0.0242 | 0.4666 |
| 207895920065_R07C01 | 0.0192 | 0.1188 | 0.0683 | 0.0555 | 0.0242 | 0.4666 |
| 207895920065_R08C01 | 0.0192 | 0.1193 | 0.0686 | 0.0557 | 0.024 | 0.4664 |
| 207895920099_R01C01 | 0.0187 | 0.1201 | 0.0689 | 0.0557 | 0.0237 | 0.4656 |
| 207895920099_R02C01 | 0.0189 | 0.1198 | 0.0688 | 0.0555 | 0.0239 | 0.4662 |
| 207895920099_R03C01 | 0.0192 | 0.1189 | 0.0682 | 0.0555 | 0.0242 | 0.4667 |
| 207895920099_R04C01 | 0.0192 | 0.1188 | 0.0682 | 0.0555 | 0.0242 | 0.4667 |
| 207895920099_R05C01 | 0.0193 | 0.1187 | 0.0682 | 0.0555 | 0.0242 | 0.4667 |
| 207895920099_R06C01 | 0.0193 | 0.1187 | 0.0682 | 0.0554 | 0.0243 | 0.4666 |
| 207895920099_R07C01 | 0.0192 | 0.1187 | 0.0682 | 0.0554 | 0.0243 | 0.4666 |
| 207895920099_R08C01 | 0.0192 | 0.1188 | 0.0683 | 0.0555 | 0.0243 | 0.4664 |
| 208553420050_R01C01 | 0.0193 | 0.1189 | 0.0683 | 0.0555 | 0.0242 | 0.4664 |
| 208553420050_R02C01 | 0.0191 | 0.119 | 0.0683 | 0.0555 | 0.0242 | 0.4663 |
| 208553420050_R03C01 | 0.0192 | 0.1189 | 0.0683 | 0.0555 | 0.0242 | 0.4664 |
| 208553420050_R04C01 | 0.0192 | 0.1189 | 0.0684 | 0.0554 | 0.0242 | 0.4664 |
| 208553420050_R05C01 | 0.0192 | 0.1189 | 0.0683 | 0.0555 | 0.0242 | 0.4664 |
| 208553420050_R06C01 | 0.0192 | 0.1188 | 0.0682 | 0.0555 | 0.0243 | 0.4665 |
| 208553420050_R07C01 | 0.0191 | 0.119 | 0.0684 | 0.0554 | 0.0243 | 0.4664 |
| 208553420050_R08C01 | 0.019 | 0.1192 | 0.0686 | 0.0555 | 0.024 | 0.4662 |
| 208553420085_R01C01 | 0.0191 | 0.1193 | 0.0683 | 0.0556 | 0.0242 | 0.4666 |
| 208553420085_R02C01 | 0.0192 | 0.1188 | 0.0682 | 0.0555 | 0.0242 | 0.4665 |
| 208553420085_R03C01 | 0.0193 | 0.1187 | 0.0682 | 0.0554 | 0.0243 | 0.4664 |
| 208553420085_R04C01 | 0.0192 | 0.1188 | 0.0683 | 0.0555 | 0.0242 | 0.4665 |
| 208553420085_R05C01 | 0.0191 | 0.1189 | 0.0683 | 0.0555 | 0.0242 | 0.4664 |
| 208553420085_R06C01 | 0.0191 | 0.119 | 0.0683 | 0.0555 | 0.0242 | 0.4664 |
